# Measuring the economic burden of breast cancer in middle-income countries: Protocol for a prospective cohort study in India and Kenya

**DOI:** 10.64898/2026.08.14.26360436

**Authors:** Beverley M. Essue, Sourajit Parida, Mansoor Saleh, Amina Kidee Habib, Debasmita Nayak, Katu Mutungi, Mitchele Midega, Linda Muriithi, Isabel Arruda-Caycho, Luca Bernardini, Maansi Kashyap, Danielle Rodin

**Author notes:** Contributed equally as second authors (Sourajit Parida, Mansoor Saleh).

## Abstract

**Background:** Gaps in health financing drive delayed diagnosis, catastrophic health expenditure, treatment discontinuation, and excess mortality and morbidity in breast cancer, effects compounded by gender inequalities that shape household resource allocation, care-seeking behaviour, and spending decisions for conditions disproportionately affecting women. Despite this, the economic burden of breast cancer and the gender dynamics that mediate it remain poorly characterised in middle-income country settings. This study examines the economic burden of breast cancer in India and Kenya and assesses how gender inequalities shape treatment decision-making, health outcomes, and caregiving experiences.

**Methods:** This will be a mixed-methods, longitudinal, prospective cohort study of newly diagnosed breast cancer patients, with a health economics and gender analysis. Participants will be surveyed twice, at baseline and 6-months post treatment commencement either in person or by phone. A sub-sample of participants and their caregivers will participate in semi-structured interviews to explore household economic consequences of treatment, treatment-seeking decisions, and the gendered dimensions of both. Quantitative data will be analysed using descriptive statistics and regression modelling to identify determinants of catastrophic health expenditure and economic burden. Thematic analysis will be conducted and triangulated with quantitative findings to provide a comprehensive account of financial and gendered impacts across both settings.

**Discussion:** The study will generate comparative evidence on the economic burden of breast cancer across two developing health system contexts. Findings will inform priority setting and benefit package design by identifying the drivers of economic burden and treatment discontinuation in these contexts, and making visible the household and caregiving costs that financing policy rarely captures.

## Background

Breast cancer is a major global health concern. It is the most common cancer in women and the most prevalent cancer worldwide [1]. It accounts for the most disability-adjusted life years in women compared to other cancers [2] and is the leading cause of cancer death in women globally [3]. When breast cancer is detected and treated early, it is associated with excellent long-term outcomes [4]. In high-income countries, 5-year survival exceeds 90 percent [5,6]. However, progress in improving survival rates has been considerably slower in most low- and middle-income countries (LMICs) [7]. Thus, breast cancer represents an ongoing challenge for global health systems and the achievement of gender equity and health equity goals [8].

In India, 5-year survival from breast cancer is still just 66.4 percent [9], with women who are poor, less educated, and live rurally at the highest risk of dying [9,10]. In Sub-Saharan Africa, the 5-year survival is estimated to be between 40 to 60 percent, influenced by stage of diagnosis, access to healthcare, availability of treatment, and public awareness [11]. Access to effective early diagnostic and treatment options remains limited for many women diagnosed with breast cancer in India and Kenya [9,12], as in other LMICs, reinforcing concerns about the widening of inequities in cancer treatment and outcomes within and between populations [8,13,14]. Ensuring equitable financing and access to breast cancer treatment is critical to improving cancer outcomes and strengthening global health systems [15].

In India and Kenya, inequitable health financing mechanisms and a mix of incomplete insurance programs have resulted in wide gaps in the coverage of breast cancer treatments, requiring patients to cover partial or full treatment costs in most settings [15–20]. About 84.6 percent of Indian households face catastrophic levels of health expenditure, which occurs when a large percent of household income or resources is spent on unreimbursed medical and health-related costs [18]. In a study of cancer patients in India, one-third of households impacted by a cancer diagnosis of a family member spent more than half of their per-capita annual household expenditure on hospitalisations [21]. In Kenya, the cost of breast cancer screening and treatment is unaffordable for the majority of women or households [17]. Unaffordable treatment costs create financial barriers to accessing and completing treatment, which can worsen cancer outcomes. Discontinuing cancer treatment due to high costs is an issue for cancer patients in India and Kenya [22–25], as well as in other LMICs [17], with patients from lower socioeconomic households facing an increased risk [17,22]. Further, social and cultural norms, including pervasive gender inequalities within societies, influence household resource allocation and decision-making, as well as health care-seeking and spending behaviours for conditions that affect women and girls [8,26,27]. Thus, the ability and willingness to pay for breast cancer treatment in LMICs can deepen inequities in access to treatment and in survival, which can be mitigated by reducing out-of-pocket costs through enhanced financial protection [18,25,28], a central goal of the Universal Health Coverage (UHC) agenda and UN Sustainable Development Goals (SDG) [29].

Relative to other chronic conditions in developing health system contexts, cancer treatment is expensive and often unaffordable for the majority of the population in need [27,28,30]. Treatment occurs over an extended period, imposes lifestyle restrictions, and is accompanied by side effects and caregiving demands [31,32], while also potentially diminishing the patient’s ability to maintain existing care responsibilities [33,34]. Ensuring financial protection entails protecting households from negative economic consequences, including impoverishment, that can result from paying for health care expenses when a household member is unwell. In the case of breast cancer, the economic burden associated with out-of-pocket costs can reverberate across families and communities, causing long-term financial consequences, and both causing and exacerbating existing impoverishment [35]. To meet treatment costs, households may deplete savings, sell assets, and borrow funds [36] at a time when household income may also be impacted [37].

Patients must also decrease their own caregiving and unpaid responsibilities in the household and community, which are predominantly carried out by women [8,33,38,39]. Informal unpaid family caregivers are relied upon to provide supportive care and consequently experience direct caregiving costs, time costs, and lost productivity [23,40]. Caregiving costs are not routinely captured in estimates of cancer-related out-of-pocket costs [41,42], but can be substantial [42,43] and can also contribute to negative economic consequences for households [44].

Further, the economic burden of a breast cancer diagnosis intersects with gender norms and roles, as breast cancer predominantly affects women and informal caregiving is predominantly carried out by females [45,46]. While gender dimensions have been shown to influence household decision-making [8,26,27], they are not routinely accounted for in the processes used to set priorities in health systems [42]. The literature on the economic burden associated with cancer in LMICs, including India and Kenya, is limited but growing, with a nascent evidence base, largely dominated by single-centre cross-sectional quantitative designs [24,47–49]. Few studies capture caregiving costs, and rarely any explicitly investigate gender dimensions [24,47,50].

## Aims and Objectives

The study will aim to centre the economic burden and financial barriers impacting breast cancer care in India and Kenya. This study will be guided by two primary objectives:

### Objective 1

To quantify and measure the out-of-pocket cost burden for individuals with breast cancer in India and Kenya and their caregivers.

### Objective 2

To qualitatively explore patient and caregiver perspectives on the economic burden of breast cancer and assess gender dimensions related to caregiving, time use, productivity losses, treatment decision-making and outcomes.

## Methods

### Overarching framework

This study will be informed by the McIntyre Access Framework [51]. The conceptualisation of the economic burden of breast cancer care will be examined across the framework’s three dimensions of affordability, availability, and acceptability, and situates out-of-pocket costs, caregiving demands, and treatment-seeking decisions within the broader structural and gendered factors that shape access to care. Through this lens, access will be understood as a multidimensional construct that characterises interactions between health care systems and individuals, and explains how these factors, like gender, systematically shape healthcare-seeking behaviour across different settings. Operationally, the framework will inform the selection of variables captured in the study instrument, including economic, clinical, sociodemographic, and gender-related domains; the structure of the qualitative interview guide; and the analytical approach.

### Study design and setting

This study will use a longitudinal, multi-methods design (Fig 1). A prospective cohort of newly diagnosed breast cancer patients will be recruited and surveyed at first presentation and at 6 months follow-up about out-of-pocket costs and the economic burden of cancer treatment and caregiving needs. Previous research suggests that costs associated with breast cancer care are typically the highest within the first 6 months of cancer diagnosis and in the final year of life [52,53]. A subsample of participants and their caregivers will be invited to participate in semi-structured interviews exploring the household economic consequences of cancer treatment and the gender impacts of treatment-seeking decisions and behaviours.

**Fig 1.**
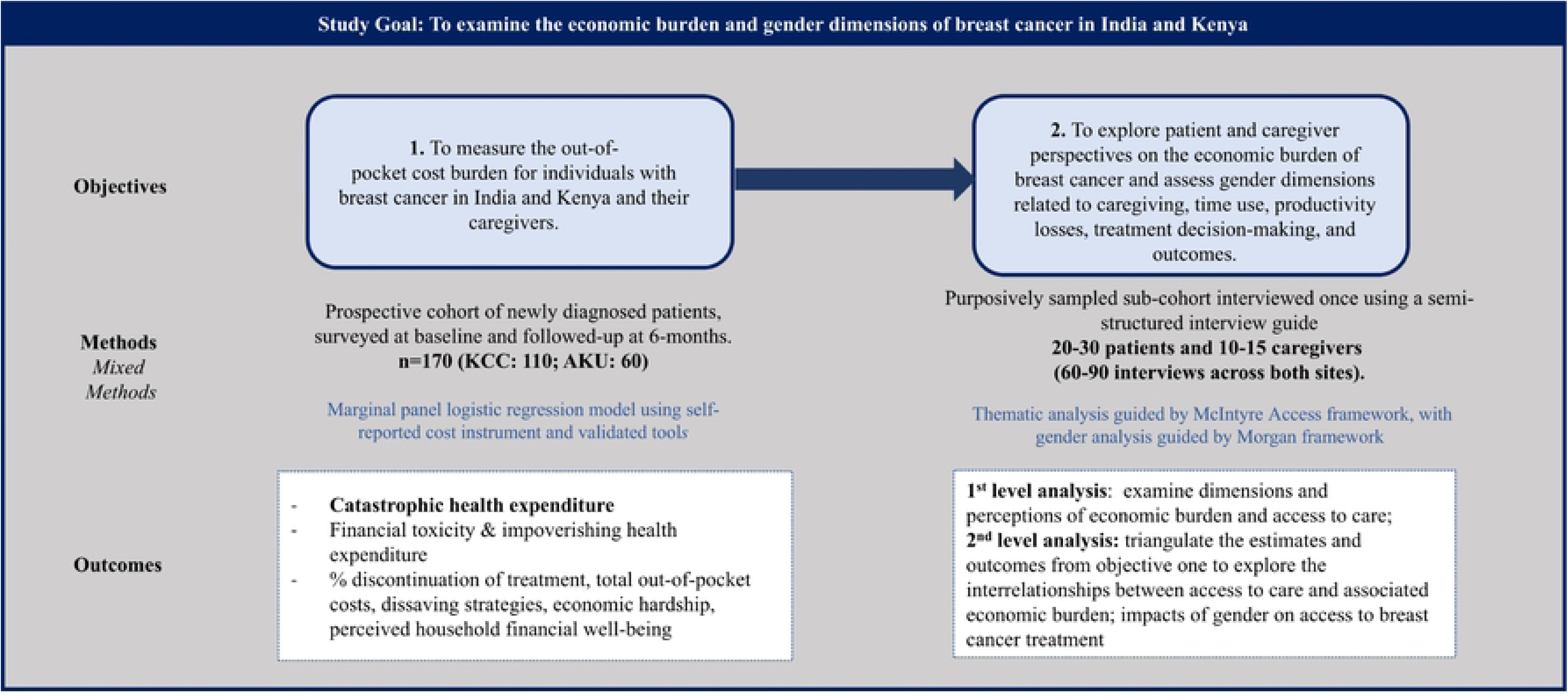
Study Methods Overview.

Data collection will occur at two cancer centres: Kalinga Institute of Medical Sciences Cancer Centre (KCC) in Bhubaneswar, India and Aga Khan University Hospital in Nairobi (AKUHN). Both centres offer cancer care management including medical, surgical and radiation oncology, as well as pain and palliative care, to cancer patients from their state and neighbouring regions. Of the new breast cancer patients seen at KCC annually, about 60% have access to private health insurance, 50% are covered under national/state health insurance schemes, and approximately 40% do not have any insurance. Of the new breast cancer patients seen at AKUHN, 65% have private health insurance, 25% have national coverage, and 10% have no insurance. In both contexts, patients with health coverage still incur some out-of-pocket treatment and health-related costs.

Both study sites are private, comprehensive cancer centres. This study’s deliberate focus on such centres will reflect the reality of how breast cancer care is predominantly financed and delivered in India and Kenya [54,55]. In both countries, the private sector accounts for a substantial share of cancer diagnosis and treatment [56,57], particularly for breast cancer [58,59], where patients frequently bypass under-resourced public facilities to access specialist oncology services [54,60,61], imaging, and systemic therapy [54,55]. Focusing on private centres will allow the examination of financial burden in the setting where much of the care occurs, and to document the gap between nominal insurance coverage and full financial protection, a gap that is poorly captured in studies conducted exclusively in public facilities where patient populations, cost structures, and insurance dynamics differ substantially [21,48,62].

### Participants and recruitment

Participants will be eligible for the study if they are cisgender females aged 18 years or older and newly diagnosed with non-metastatic breast cancer in the past three months. Additionally, participants must be aware of their diagnosis and must be cognitively able to provide consent and to participate in data collection.

Participants must be accrued to the study within 12 weeks of first presentation at the clinic following breast cancer diagnosis. Participants with a synchronous or prior history of cancer (breast cancer or other cancer) will be excluded from the study. Cisgender men and transwomen will be excluded because a sufficient sample size is not anticipated to estimate sex-specific outcomes due to the low incidence of breast cancer among individuals assigned male at birth [63,64], and because the study will focus on how the female contextual gender social constructs shape access and economic burden.

Eligibility will be determined by the site investigator through a histologically confirmed diagnosis of breast cancer, as indicated in medical records and investigations. Participants with a confirmed new diagnosis who meet the inclusion criteria will be invited to participate in the study by a research assistant based in each hospital site. Participant information sheets and consent forms will be provided to eligible participants and explained by a member of the research team. Participants will have the opportunity to discuss the study and ask questions prior to providing informed consent. Participants will be recruited at KCC over 18 months and at AKU over 12 months to achieve the required sample size.

The sample size will be calculated using the probability-proportional-to-the-size-method [65]. KCC treats approximately 200 new breast cancer patients each year and AKU treats approximately 144 new breast cancer patients each year (all stages) [66]. Based on a previous single-state study that estimated 84% of breast cancer patients experienced catastrophic health expenditure (CHE), for 5% alpha and assuming 20% attrition, 110 participants from KCC and 60 participants from AKU will be recruited for a study sample of 170 participants.

### Data collection and instrument

#### Objective 1

Baseline data collection will occur within 12 weeks of first presentation and will be conducted in-person during participants’ scheduled hospital appointments by a research team member based at each hospital. Follow-up will occur 6 months after the baseline data collection point, either in-person during a hospital visit or by phone. Where death occurs, the participant’s named alternative respondent will complete the follow-up. Honoraria will be offered to participants as compensation for their time and reimbursement for any research-related costs.

Currently, no existing validated tools comprehensively capture the costs and economic burden associated with breast cancer for use in LMICs. For the collection of information on out-of-pocket costs and the measurement of the economic burden of cancer care, self-reported instruments are the preferred method [20,23,67]. The self-reported study instrument will be adapted from tools previously used by the research team and translated into Odia and Kiswahili, the two primary languages spoken in the study settings. A paper-based copy of the instrument will be administered by a trained research assistant at each site.

The study instrument will consist of a questionnaire administered at baseline and a modified questionnaire administered at follow-up to collect vital status and verify if patients are alive or deceased (S1 Table). Deceased status will be confirmed using medical records or by a named caregiver. The baseline questionnaire will collect individual, household, clinical and system-level information, including socio-demographic information, medical history, cancer treatment plan and intent, household finances and income, healthcare utilization, and caregiver and gender-related variables. The translated versions of the instrument will be piloted with 10% of the target sample at each site.

Standardized measures that have been validated for assessments with cancer populations and in healthcare settings will be used to collect information on financial toxicity, mental health, and quality of life. The study will assess financial toxicity with version 2 of the COmprehensive Score for Financial Toxicity–Functional Assessment of Chronic Illness Therapy tool (COST-FACIT-v2) [68,69], a validated patient-reported outcome tool designed to assess financial toxicity among cancer patients, that will be validated in Odia [70] and Kiswahili to support application in Indian and Kenyan contexts. Quality of life will be assessed using version 3 of the European Organization for Research and Treatment of Cancer Core Quality of Life Questionnaire (EORTC QLQ-C30) [71], and mental health will be assessed using the Depression, Anxiety and Stress Scale–21 (DASS-21) [72].

The primary outcome for this study will be catastrophic health expenditure (CHE), a key indicator for monitoring progress toward the UHC agenda and the United Nations SDGs on financial protection [73–75]. CHE will be defined as the proportion of households with annual out-of-pocket expenditures exceeding 10% of income [76]. The secondary outcomes will be financial toxicity (i.e., mean standardized score from the COST-FACIT-v2 instrument) and impoverishing health expenditure (i.e., proportion of households in which household income minus annual out-of-pocket costs is below the national poverty line), recognizing that these secondary outcomes are conceptually distinct from CHE and may have different predictors. We will use the Kenya Integrated Household Budget Survey and Tendulkar Poverty Line to define the inflation-adjusted 2024 national urban poverty lines in Kenya and India, respectively [77,78]. A sensitivity analysis will be conducted using the World Bank Lower Middle Income Class poverty line as an alternative threshold for defining impoverishing health expenditure [77,79].

Additional outcomes to be reported descriptively are: annual out-of-pocket costs (i.e., mean out-of-pocket costs extrapolated to 12 months); use of dissaving strategies (e.g., selling assets or borrowing money); economic hardship (i.e., responding “yes” to missing any of the specified payments); perceived household financial well-being (i.e., the proportion of patients reporting each category of self-rated financial prosperity), and cost-related treatment abandonment (i.e., the proportion of patients who respond “yes” to questions about treatment discontinuation during follow-up data collection). Changes in outcomes between baseline and follow-up will be assessed using mean changes for continuous variables and changes in proportions for categorical variables.

#### Objective 2

A sub-cohort of patients and caregivers will be purposively sampled according to key variables to maximize sample heterogeneity and variation across age, cancer stage, insurance type and income [80], and invited to complete 30-minute qualitative interviews separately and linked as a dyad in the analysis. Participants will be recruited until saturation is achieved. Saturation is anticipated to occur after approximately 20 to 30 patient interviews and 10 to 15 caregiver interviews at each site.

Semi-structured interview guides will be developed and pilot tested with 2 patient-caregiver dyads in each setting, approximately 10% of the target sample size for each study group. Interviews will be conducted in-person or by phone [81] by a trained research assistant. Interviews will be conducted in English, Kiswahili, or Odia, and audio-recorded to generate an interview transcript that will be translated to English for subsequent analysis. The qualitative interview guide will be structured around the three dimensions of the Access Framework to ensure that participants’ accounts of treatment-seeking decisions, household expenses, and gendered experiences of care are systematically explored in relation to the broader health system and social factors that shape access (S1 Fig). The interviews will examine the impact of the cancer diagnosis on household economics, perceptions of the availability, acceptability, and affordability of cancer treatment, strategies to pay for cancer treatment and their impacts, and household decision-making, including gender constructs that impact patient involvement in decision-making about treatment [83]. The caregiver interviews will explore cancer-related caregiving demands and impact on their health, quality of life, satisfaction with role, as well as gender constructs associated with caregiving expectations.

## Data Management

Study sites will complete data collection on deidentified paper copies of the study instrument and will subsequently enter the data into a shared Research Electronic Data Capture (REDCap) database to store for analysis. Paper copies and any identifying information, including participant names and hospital numbers, will be securely stored at each hospital site and will remain separate from study data.

Routine REDCap data quality checks on 15% of available data per study site will be implemented and shared during quarterly across-site team meetings. Quality checks will focus on examining records for field completeness to identify missing or invalid data entries, as well as ensuring REDCap functionality. Source verification will be completed by research assistants to confirm concordance between REDCap records and paper-based copies.

## Data Analysis

### Objective 1

First, out-of-pocket cost data are expected to contain a substantial proportion of zero values and are unlikely to satisfy the assumption of normally distributed errors. A two-part model will therefore be used. A logistic regression using Generalized Estimating Equations (GEE) will estimate the probability of incurring any OOP health expenditure, and a generalised linear model (GLM) estimated using GEEs will estimate the level of expenditure among those with positive costs [82,83]. This approach is well-suited to the skewed, zero-inflated distributions typical of health care cost data in LMIC settings. With the two-part model, we will first identify who incurs any OOP expenditure before we examine who experiences catastrophic health expenditure (CHE), impoverishing health expenditure, and financial toxicity through regression models. All regression analyses will be two-sided with a significance level of 0.05.

The primary outcome, CHE, will be measured as a binary variable, coded as 1 if a household’s out-of-pocket (OOP) health payments exceed the defined threshold and 0 otherwise. The two secondary outcomes, impoverishing health expenditure and financial toxicity, will also be measured as binary variables. Impoverishing health expenditure will be coded as 1 if household income minus out-of-pocket (OOP) health payments falls below the national poverty line, and 0 otherwise. Financial toxicity will be coded as 1 if the respondent’s COST-FACIT-v2 score falls below 26, and 0 otherwise, according to established thresholds reported in the literature for the original COST-FACIT instrument [84]. The same threshold will be applied to version 2 of the instrument because the metric and underlying construct for each item are consistent across versions. McNemar’s tests will be used to assess the change in the incidence of primary and secondary outcomes between baseline and follow-up. Marginal panel logistic regression models, implemented with Generalized Estimating Equations (GEE), will be used to estimate the population-level predictors of the primary and secondary outcomes, accounting for within-household correlation over time. Variables will be selected for regression analysis using a combination of statistical significance in bivariate analysis (p < 0.05) and a priori selection guided by Andersen’s Health Care Utilization Model and existing literature [85].

To examine the additional outcomes: Expenditure data collected at baseline and six-month follow-up will be combined to estimate annual out-of-pocket costs, which will be reported in aggregate and disaggregated into direct medical costs and broader health-related costs, including transport, accommodation, and caregiving expenditures [66]. The use of dissaving strategies, economic hardship, perceived financial well-being, and cost-related treatment abandonment will each be assessed using paired t-tests or Wilcoxon signed-rank tests, as appropriate, and mixed-effects regression models.

An equity analysis will be directly informed by the affordability dimension of the Access framework, which positions out-of-pocket costs not as individual financial events, but as systemic consequences of how care is financed and organised. To characterize the distributional consequences of out-of-pocket costs and CHE, outcomes will be stratified by household income bands to assess how these outcomes are distributed across income groups and to descriptively identify sub-populations bearing disproportionate risk of CHE and high out-of-pocket costs.

All analyses will be conducted separately for each site and results will be reported independently to reflect the distinct health system contexts, insurance systems, and cost structures in India and Kenya. Pooled analyses will be conducted where appropriate with site included as a fixed effect. In pooled regression models, site will be entered as a binary indicator variable, with one site designated as the reference category, allowing estimation of a common effect of covariates on outcomes while also accounting for systematic between-site differences in the baseline level of the outcomes. Where sample sizes permit, subgroup analyses will be conducted by insurance status, disease stage, and treatment modality. Costs will be expressed in 2025 local currency and converted to international dollars using purchasing power parity conversion factors to support cross-country and cross-study comparison. All analyses will be conducted using R.

### Qualitative

Qualitative thematic data analysis will follow an interpretivist paradigm [86]. Interview transcripts will be examined to identify concepts or ideas that relate to economic burden and access to care to develop themes. Python Programming will be leveraged to support the development of a study codebook and Natural Language Processing (NLP) will be applied to identify frequently present codes and topic modelling. Codes and topics will be interpreted by the research team into overarching themes. While the initial study proposed, written several years ago, planned for manual coding, as Python Programming and generative AI were less commonly used to support qualitative data analyses, the plan was revised to incorporate these contemporary tools and programming language now more widely used in qualitative research. A narrative will be developed to elucidate each of the themes, which is essential for validating assumptions in the data interpretation process [87].

The second level of analysis will utilize Python Programming to triangulate the estimates and outcomes from objective one with the qualitative findings to explore the interrelationships between access to cancer treatment and associated economic burden. The study will implement a gender analysis utilizing the Morgan framework [88] to investigate gender as a power relation and driver of inequality in access to breast cancer treatment. A sub-sample of study participants and the leadership team will be engaged to support and validate the interpretation of the findings.

### Ethical Considerations

Ethics approval will be sought and obtained from the Aga Khan University, Nairobi Institutional Scientific and Ethics Review Committee (ISERC) [2024-ISERC-172 (v2)], the Kalinga Institute of Medical Sciences Institutional Ethics Committee [1844/2024], and University of Toronto Research Ethics Board [44562] prior to recruitment initiation. Verbal and written informed consent will be sought from all participants by a research team, and they will be provided with site investigator and ethics board contact information. They will be informed that they are free to withdraw from the study at any time without affecting their care or their relationship with the hospital site. Participants will be asked to share the contact details of a caregiver or an alternative respondent who could complete the interview in case of participant demise, in which case the additional consent of the alternative respondent will be obtained.

### Anticipated Timeline

Recruitment and data collection began on April 01, 2025. We anticipate all recruitment, data collection, and cleaning will be completed by December 2026. Results and manuscript publication are expected by early 2027. The site-specific research teams meet fortnightly and the full study team meetings are scheduled quarterly.

## Discussion

A few methodological challenges are anticipated. The self-report survey and interview design give way to potential research biases, including response and interviewer biases, which will be mitigated by crafting reflexive interview questions, providing adequate training for research assistants, and utilizing validated instruments and questions previously used in other published studies. To prevent sampling bias in the qualitative interviews, purposive sampling across various socio-demographic characteristics, including age, cancer stage, income group and insurance type, will be used in the qualitative element of this study to prevent potential sampling biases. Additionally, participant mobility may complicate follow-up, and sustaining coordination across a large international team introduces logistical complexity. These risks are mitigated by a prospective design with conservative power calculations that account for anticipated attrition, mobile phone follow-up to reduce participant burden, honoraria to reimburse participation costs, periodic data quality checks, and regular synchronous research team meetings. The feasibility of recruiting and following a comparable cohort of incident breast cancer patients in similar settings has been demonstrated by members of the investigator team in prior work, providing confidence in the achievability of the proposed recruitment targets.

The study will have several important strengths. It is anchored in genuine co-designed, equitable research partnerships with established cancer centres in India and Kenya, both of which have active research programs with demonstrated capacity to recruit and follow cancer patients and caregivers. The investigator team will bring complementary expertise in cancer treatment cost research, health financing, clinical oncology, and gender-responsive research design across LMIC settings. The integration of validated, context-specific instruments, including the COST-FACIT-V2, translated and validated in Odia and Kiswahili for this study, strengthens measurement rigour and cross-country comparability. The mixed-methods design, combining prospective cost data with in-depth qualitative inquiry into treatment-seeking decisions and household economic responses, will enable a more complete account of financial burden than quantitative measurement alone can provide.

Critically, the study is designed not only to document the existence of economic burden but to characterise its distribution, identify its drivers, and make visible the gendered dimensions of household economic decision-making that standard cost analyses routinely overlook. These are precisely the dimensions of evidence needed to move financial protection for cancer from a measurement problem to a policy design problem. By generating equity-stratified cost data anchored in the care pathways of two complementary settings, the study will produce findings directly applicable to benefit package design, priority-setting, and advocacy for expanded cancer coverage in India and Kenya, with relevance to a broad range of LMIC contexts where breast cancer burden is growing and financial protection remains inadequate.

Findings from this study will be developed into an academic manuscript and submitted to a peer-review journal. A lay summary will be developed and shared with participants and health professionals. In summary, this protocol describes a prospective cohort study examining the economic burden of breast cancer and its gendered dimensions across two middle-income country settings with distinct health system architectures. The study addresses a well-documented gap in the literature: while financial hardship from cancer is known to be substantial in LMICs, evidence on its drivers, its distribution across income groups, and its gendered dimensions remains limited, particularly in sub-Saharan Africa and South Asia where the burden is rising fastest.

## Data Availability

Deidentified data from this study can be requested from the authors upon study completion.

## Author’s Contributions

BME, DR conceived of the study, led the funding application and provided oversight and leadership for the research. SP, AKH, MS provided clinical and scientific expertise to refine the project design and contextualize it for both study contexts. IC-A supported overall study management, including overseeing data management and quality. DN, MM, LM, conducted all data collection and coordinated the research at each site. LB and MK provided research assistance, including supporting the translation of tools and the development of the analysis plan. All authors contributed to drafting this protocol manuscript and reviewed and approved the final draft.

## Acknowledgements

The authors are grateful to Bidhu Mohanti and Sujata Mishra for their support in developing the funding proposal and in facilitating the early discussions that enabled the initiation of the study at KCC. The authors also thank the advisory committee for their support during the conceptualization of the project.

## Supporting Information

**S1 Table: Overview of Data Elements, Measures and Data Collection Schedule.**

**S1 Fig: Overview of Data Elements Investigated in Qualitative Interviews.**

